# Healthcare personnel knowledge, motivations, concerns and intentions regarding COVID-19 vaccines: a cross-sectional survey

**DOI:** 10.1101/2021.02.19.21251993

**Authors:** Vivek Jain, Sarah B. Doernberg, Marisa Holubar, Beatrice Huang, Carina Marquez, Lillian Brown, Luis Rubio, Hannah A. Sample, Jenna Bollyky, Guntas Padda, Daisy Valdivieso, Amanda Kempema, Christopher Leung, Matthew Sklar, Aida Julien, Marcus Paoletti, Sravya Jaladanki, Emerald Wan, Jacob Ghahremani, Jessica Chao, Yingjie Weng, Di Lu, David Glidden, Kevin Grumbach, Yvonne Maldonado, George Rutherford

## Abstract

**Background:** Healthcare personnel (HCP) are prioritized for earliest SARS-CoV-2 vaccine administration, yet relatively few data exist on HCP’s knowledge, motivations, concerns, and intentions regarding COVID-19 vaccines.

**Methods:** We conducted a cross-sectional survey Nov.16-Dec.8, 2020 among HCP enrolled in a cohort study at three Northern California medical centers serving diverse roles including COVID-19 patient care. Eligible HCP were adult (age≥18) on-site employees of the University of California, San Francisco, San Francisco General Hospital, and Stanford Healthcare. A one-time electronically-administered survey was sent to cohort HCP on November 16, 2020 and responses analyzed.

**Results:** Overall, among 2,448 HCP invited, 2,135 completed the COVID-19 vaccine survey (87.2% response rate). HCPs had mean age 41 years, were 73% female, and had diverse jobs including COVID-19 patient contact. Enthusiasm for vaccination was overall strong, and more HCP (1,453, 69%) said they would definitely/likely receive vaccine if formally FDA-approved versus if approved via emergency use authorization only (785, 35%). While 541 (25%) respondents wanted to be among the earliest to receive vaccine, more desired vaccination after the first round (777, 36%) or >2 months after vaccinations began (389, 18%). Top factors increasing motivation for vaccination included perceiving risk from COVID-19 to self (1,382, 65%) or to family/friends (1355, 63%). Top concerns were vaccine side effects, cited by 596 (28%), and concerns about political involvement in FDA’s approval process (249, 12%).

**Conclusions:** HCP were enthusiastic about COVID-19 vaccination for individual protection and protecting others, but harbored concerns about vaccine side effects. Our data may inform emerging vaccine education campaigns.

**Key Points:** Among 2,135 healthcare personnel surveyed, we found enthusiasm for COVID-19 vaccination both for individual benefit and protecting others. However, healthcare personnel rated their knowledge of COVID-19 vaccines as only moderate and harbored concerns about vaccine side effects. Education raising awareness of vaccine efficacy and side effects may help maximize vaccine uptake.

## Introduction

Two mRNA vaccines against SARS-CoV-2 made by Pfizer/BioNTech [1] and Moderna [2] recently showed 95% and 94.1% efficacy and received U.S. Food and Drug Administration (FDA) emergency use authorization (EUA) in December 2020. The Centers for Disease Control (CDC) Advisory Committee on Immunization Practices endorsed both COVID-19 vaccines [3, 4] and designated healthcare personnel as top priority for immediate vaccination. [5] However, relatively little is known about healthcare personnel’s knowledge, attitudes, motivations and intentions surrounding COVID-19 vaccines. To address these questions and inform emerging vaccine education campaigns, we surveyed frontline healthcare personnel at three academic medical centers about COVID-19 vaccines.

## Methods

### Study setting

The CHART Study (COVID-19 Healthcare Worker Antibody and RT-PCR Study) is a longitudinal cohort study of adult (age≥18) healthcare personnel working at the University of California, San Francisco (UCSF), Zuckerberg San Francisco General Hospital, and Stanford Health Care during the pandemic. Cohort members enrolled from July-November 2020 at all three centers. Persons from all job types were eligible to enroll, regardless of whether they directly cared for patients with COVID-19. Exclusion criteria included not working on site during the pandemic or planning to end employment in <6 months.

### Survey

Participants were sent an electronic link to a web-based eight-question web-based survey via REDCap (Research Electronic Data Capture) about COVID-19 knowledge, motivations, concerns, and intentions on November 18, 2020. Participants completed surveys over three weeks. Data were analyzed on December 8, 2020.

### Analysis

Proportions of respondents answering each question were tabulated and analyzed.

### Ethics

The CHART Study and the COVID-19 vaccine survey were approved by the UCSF Committee on Human Subjects Research and the Stanford University School of Medicine Panel on Human Subjects in Medical Research.

### Patient Consent Statement

All participants provided informed written consent for participation.

## Results

Of 2,135/2,448 (87.2%) healthcare personnel completed the COVID-19 vaccine survey. Healthcare personnel had mean age of 41.4 years and were 73% female (Table 1). Overall, 839 (39.3%) were registered nurses, and 478 (22.4%) were physicians. Work locations included intensive care units (11.5%), hospital wards (17.8%), and emergency department/urgent care (7.9%). In total, 574 (26.9%) provided direct patient care involving airway intubation/suctioning, and 1,050 (49.2%) provided direct patient care not involving airway procedures.

**Table 1.** Demographics and work characteristics of 2,135 healthcare worker respondents.

|  | Healthcare workers<br>(n=2135) |
| --- | --- |
| **Mean age, years (SD)** | 41.4 (10.2) |
| **Gender, n (%)** |  |
| Male | 396 (18.5) |
| Female | 1563 (73.2) |
| Other | 10 (0.5) |
| Unknown/Missing | 166 (7.8) |
| **Race/Ethnicity, n (%)** |  |
| Non-Hispanic White | 1126 (52.7) |
| Non-Hispanic Black | 26 (1.2) |
| Hispanic | 218 (10.2) |
| Non-Hispanic Asian | 426 (20.0) |
| Other<sup>A</sup> | 172 (8.1) |
| Unknown/Missing<sup>B</sup> | 167 (7.8) |
| **Healthcare Job (%)** |  |
| Attending/staff physician | 353 (16.5) |
| Registered nurse | 839 (39.3) |
| Resident/fellow physicians | 125 (5.9) |
| Advanced practitioner<sup>C</sup> | 209 (9.8) |
| Other<sup>D</sup> | 445 (20.8) |
| Missing | 164 (7.7) |
| **Area of Work (%)** |  |
| Intensive care units | 246 (11.5) |
| Wards: internal medicine | 215 (10.1) |
| Wards: other specialties | 165 (7.7) |
| Emergency department | 79 (3.7) |
| Acute/urgent/express care units | 89 (4.2) |
| Work in multiple units of hospital | 444 (20.8) |
| Other | 720 (33.7) |
| Unknown/Missing | 177 (8.3) |
| **Job Categories (%)** |  |
| Direct patient care with airway intubation/suctioning | 573 (26.8) |
| Direct patient care without any airway procedures | 1050 (49.2) |
| Indirect patient contact | 105 (4.9) |
| Laboratory technicians/staff | 49 (2.3) |
| Work not in setting with patients or biological samples | 78 (3.7) |
| Other | 116 (5.4) |
| Unknown/Missing | 164 (7.7) |

A: Includes multiple races
B: Includes individuals who declined to answer
C: Physician assistant, nurse practitioner, California registered nurse anesthetist
D: Includes food services/nutritionists (11), medical or nurse assistant or technologist (44), microbiology laboratory worker (7), pathology and morgue workers (9), registration and front desk staff (13), transport (1), phlebotomy (8), physical and occupational therapy (43), radiology technician (33), respiratory/speech therapist (22), ward clerk (4), and other (250)

On a 5-point graduated scale, only 411/2,135 (19%) respondents reported being highly informed (3%) or well informed (16%) about COVID-19 vaccine candidates (Fig.1). Most reported an average (30%) or somewhat informed (36%) level of knowledge, and 314 (16%) reported they were not informed at all.

**Figure 1.**
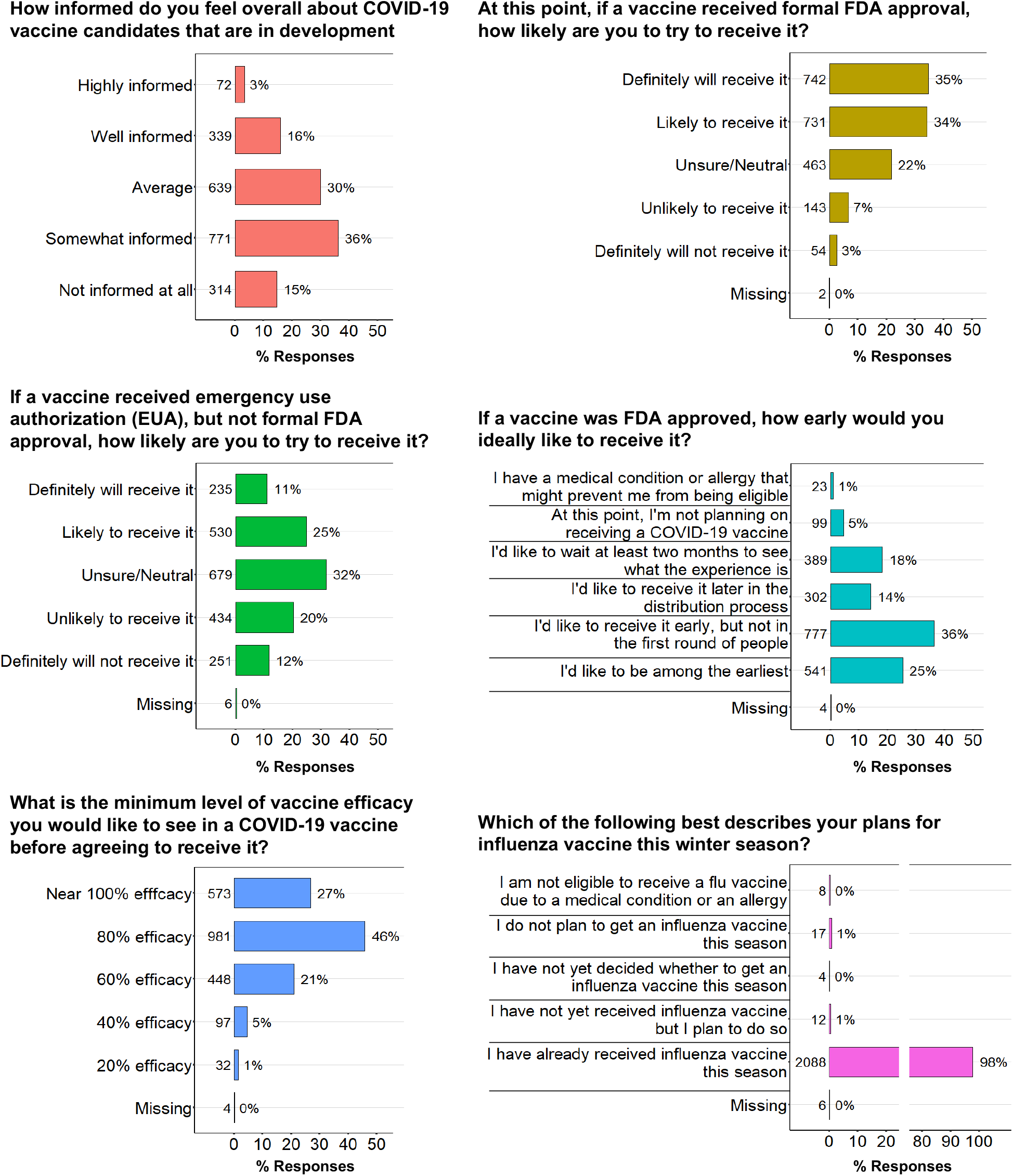
Healthcare personnel’s knowledge and opinions on COVID-19 vaccines. Numbers and proportions of respondents to individual questions on aspects of COVID-19 vaccines are shown.

When asked about intention to receive a COVID-19 vaccine with formal FDA approval, 1,453 (69%) indicated they would definitely (35%) or likely (34%) receive it. Only 10% indicated they were unlikely to or definitely would not receive it. When asked about receiving a COVID-19 vaccine that only had FDA emergency use authorization, but lacked formal FDA approval, only 36% said they would definitely (11%) or likely (25%) receive it; this was 48% lower than if vaccine was fully FDA-approved.

When asked about vaccine timing, 541 (25%) indicated desire to be among the first to receive it, while 36% desired receiving it early, but not in the first round. Conversely, 18% indicated desire to wait >2 months before vaccination, and only 5% said they were not planning to receive it at all.

In total, 573/2,135 (27%) of healthcare personnel indicated they wanted a COVID-19 vaccine to have near 100% efficacy before receiving it, and 981 (46%) desired 80% vaccine efficacy. Only 448 (21%) said they would receive a vaccine having 60% efficacy. To understand healthcare personnel’s prior vaccination patterns, we queried their status of influenza vaccination in the same winter season. The vast majority of respondents (2088 [98%]) reported already receiving influenza vaccine this season.

Finally, we queried what factors would increase motivation and desire for a COVID-19 vaccine. The highest-ranked factor was a perception of self-risk of risk of acquiring COVID-19 cited by 1,382 (65%) respondents as a strong or somewhat strong factor (Fig.2, Panel A), though only 30% perceived a high personal medical risk of COVID-19 complications. Perception of placing family members and friends at some risk of COVID-19 was cited by 1,355 (63%) as a strong or somewhat strong factor. Factors cited as mildly or not at all increasing desire for COVID-19 vaccination included a self-perception of high medical risk and COVID-19 complications (1,491, 70%), living with individuals at high risk of acquiring COVID-19 (1,442, 67%), or living with individuals at higher risk of COVID-19 complications (1,442, 67%).

**Figure 2.**
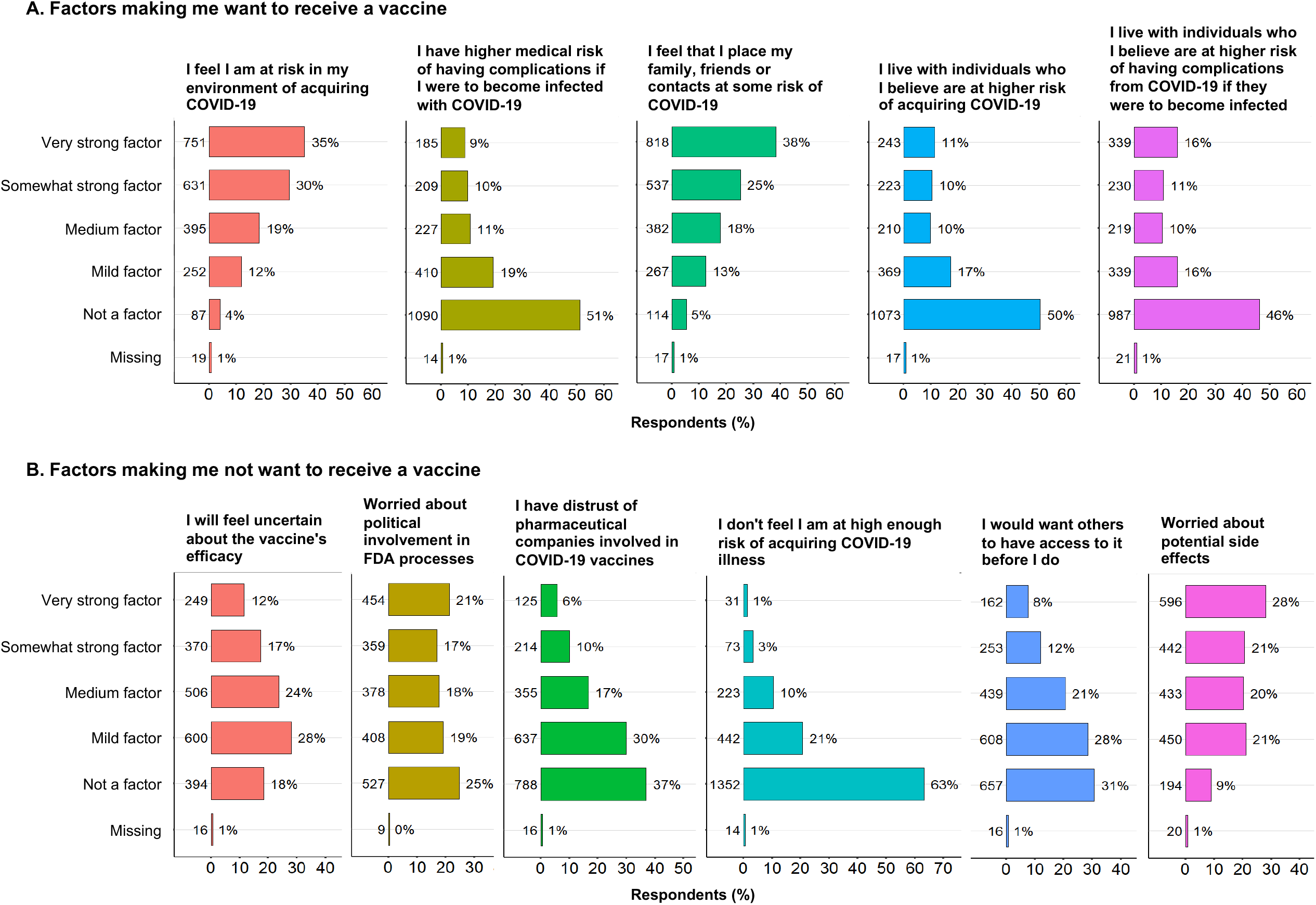
Factors influencing healthcare personnel’s opinions for and against COVID-19 vaccines. Numbers and proportions of respondents to individual questions on factors making healthcare personnel want to receive a COVID-19 vaccine (Panel A), and factors making healthcare personnel not want to receive a COVID-19 vaccine are shown (Panel B).

Top-ranked factors making healthcare personnel not want to receive COVID-19 vaccination included worry about potential side effects (596, 28%), political involvement in FDA vaccine evaluation and approval processes of vaccine evaluation and approval (454, 21%), and uncertainty about vaccine efficacy (249, 12%, [Fig.2, Panel B]). Distrust of pharmaceutical companies was only cited by 339 (16%) as a strong or somewhat strong factor decreasing desire for COVID-19 vaccination. Distrust of pharmaceutical companies was only cited by 339 (16%) as a strong or somewhat strong factor influencing them against wanting a COVID-19 vaccine.

## Discussion

On the eve of regulatory approvals of the first two COVID-19 vaccines,[6, 7] we surveyed over 2,000 healthcare personnel at three large Northern California medical centers and found overall very high enthusiasm and intention to be vaccinated for the protective benefits and secondary benefits to family and friends. However, we also found concerns about vaccine side effects and efficacy, and about political influence on regulatory approval processes. To our knowledge, this is one of the largest surveys to date of U.S. healthcare personnel on knowledge and intentions regarding COVID-19 vaccines.

Our findings may help inform public education campaigns directed at healthcare personnel and the general public to promote uptake of COVID-19 vaccines. Additionally, since many individuals will seek information about COVID-19 vaccines from healthcare providers, our data may also help reinforce reasons for vaccination and enable healthcare personnel to respond to concerns and provide reassurance where possible.[8]

Our findings on motivations for and concerns about vaccination are similar to a September survey in which Los Angeles healthcare personnel cited self-protection and preservation of community health as key reasons for COVID-19 vaccination and expressed confidence in vaccine safety and efficacy.[9] However, in that study 67% of healthcare workers cited an intention to delay COVID-19 vaccination initially, and 47% expressed reluctance about participating in a COVID-19 vaccine clinical trial, far higher than we found in our more recent survey. Likewise, in our study, we found more enthusiasm for vaccination than in a recent study conducted at SUNY Syracuse in November 2020 in which only 58% of the healthcare personnel surveyed expressed any intent to be vaccinated. [10] Notably, our population had a higher degree of racial and ethnic diversity; 53% of our study cohort are white compared with 85% of the SUNY Syracuse population surveyed. Among French healthcare personnel surveyed even earlier in the pandemic in March-July 2020, 77% intended to receive COVID-19 vaccine, motivated primarily by self-risk. [11] Notably, only 57% of respondents reported receiving influenza vaccine, far lower than the nearly 100% rate we observed.

Healthcare personnel’s motivations for COVID-19 vaccination mirror those seen in the general population. In a survey of U.S. adults in April 2020, 58% reported intent to receive a COVID-19 vaccine. [12] Factors predicting vaccine hesitancy included younger age, African American race, and not having influenza vaccine in the prior year. Notably, the surveys of the U.S. general public [12] and French healthcare personnel [11] were both done early in the pandemic, long before recent reports on vaccine efficacy and safety.

Our study contains certain limitations: it was conducted rapidly from mid-November to early December 2020 in a dynamic environment of emerging data on COVID-19 vaccines. Efficacy and side effect data on the Pfizer-BioNTech [1]and Moderna [2] vaccines were made public during this time. These data may have been available to some but not other respondents. Further, FDA decisions on the two vaccines were not yet available, and emergency authorization for Pfizer-BioNTech and Moderna vaccines occurred, respectively, on December 11 and 18, 2020 [6, 7]—after survey completion. It likely that healthcare personnel’s opinions on vaccination will evolve over time; future surveys and ascertainments of vaccination completion are planned to capture these changes.

In summary, among 2,135 Californian healthcare personnel, we found moderate knowledge about COVID-19 vaccines, strong enthusiasm and intention for vaccination, but also certain concerns about possible side effects. Our data may help inform vaccine education campaigns for both healthcare personnel and the general public.

## Data Availability

Contact corresponding author Vivek Jain at.

## Acknowledgements

Authors gratefully acknowledge the participation of healthcare personnel in the CHART Study at UCSF Health and San Francisco General Hospital of the University of California, San Francisco, CA, and at Stanford Heath Care, Palo Alto, CA.

